# The impact of heating, ventilation, and air conditioning design features on the transmission of viruses, including the 2019 novel coronavirus: a systematic review of filtration

**DOI:** 10.1101/2021.09.23.21264025

**Authors:** Gail M. Thornton, Brian A. Fleck, Emily Kroeker, Dhyey Dandnayak, Natalie Fleck, Lexuan Zhong, Lisa Hartling

**Author notes:** **Correspondence:** Dr. Brian A. Fleck; Department of Mechanical Engineering, Faculty of Engineering, University of Alberta; 116 Street and 85 Avenue; Edmonton, Alberta, Canada T6G 2R3.

## Abstract

Historically, viruses have demonstrated airborne transmission. Emerging evidence suggests the novel coronavirus (SARS-CoV-2) that causes COVID-19 may also spread by airborne transmission. This is more likely in indoor environments, particularly with poor ventilation. In the context of potential airborne transmission, a vital mitigation strategy for the built environment is heating, ventilation, and air conditioning (HVAC) systems. HVAC features could modify virus transmission potential. A systematic review following international standards was conducted to comprehensively identify and synthesize research examining the effectiveness of filters within HVAC systems in reducing virus transmission. Twenty-three relevant studies showed that: filtration was associated with decreased transmission; filters removed viruses from the air; increasing filter efficiency (efficiency of particle removal) was associated with decreased transmission, decreased infection risk, and increased viral filtration efficiency (efficiency of virus removal); increasing filter efficiency above MERV 13 was associated with limited benefit in further reduction of virus concentration and infection risk; and filters with the same efficiency rating from different companies showed variable performance. Increasing filter efficiency may mitigate virus transmission; however, improvement may be limited above MERV 13. Adapting HVAC systems to mitigate virus transmission requires a multi-factorial approach and filtration is one factor offering demonstrated potential for decreased transmission.

**Practical Implications:** In order for filtration to be an effective means of virus removal and transmission control, proper installation is required. As well, professionals should be aware of the fact that similarly rated filters from different companies may offer different virus reduction results. While increasing filtration efficiency (i.e., increasing MERV rating or moving from MERV to HEPA) is associated with virus mitigation, there appears to be diminishing returns for filters rated MERV 13 or higher. Although costs increase with filtration efficiency, filtration costs are lower than the cost of ventilation options with the equivalent reduction in transmission.

## Introduction

In March 2020, the World Health Organization (WHO) declared a global pandemic due to Coronavirus Disease 2019 (COVID-19) which is attributed to Severe Acute Respiratory Syndrome Coronavirus 2 (SARS-CoV-2).^1^ Worldwide, public health authorities have sought evidence regarding routes of virus transmission and corresponding public health measures to mitigate virus spread. Certain viruses can be transmitted via an aerosol route.^2^ Aerosol transmission is facilitated by virus-laden aerosols expelled by humans, remaining airborne for extended periods of time. Recent evidence suggests that SARS-CoV-2 can spread via airborne transmission, particularly in indoor environments with poor ventilation.^3-4^ In April 2021, the American Society of Heating, Refrigerating, and Air-Conditioning Engineers (ASHRAE) released a statement declaring that “[a]irborne transmission of SARS-CoV-2 is significant and should be controlled. Changes to building operations, including the operation of heating, ventilating, and air-conditioning systems, can reduce airborne exposures.”^5^ Determining appropriate measures to protect occupants of indoor spaces based on informed, interdisciplinary research is critical to managing the spread of infectious disease.^6^

Heating, ventilation, and air conditioning (HVAC) systems can mitigate airborne transmission of viruses by removing or diluting contaminated air inside the building envelope where humans breathe.^6-9^ HVAC design features, such as, filtration, ventilation, ultraviolet radiation, and humidity can influence transmission. The first official air filter testing standard in the HVAC industry was published by ASHRAE in 1968: ASHRAE 52-68.^10^ In 1976, the updated ASHRAE 52-76 was published. In Europe, ASHRAE 52-76 was adapted as Eurovent 4/5 which classified filters from EU1 to EU9.^10^ In 1999, ASHRAE 52.2 introduced the classification Minimum Efficiency Reporting Value (MERV). ASHRAE 52.2 was modified in 2007, 2012, and 2017. By 2016 in Europe, ISO 16890 replaced EN779 which classified filters as G1-G4; M5-M6; F7-F9.^10^ EN779 had replaced Eurovent 4/9 which superseded Eurovent 4/5.^10^ In ASHRAE Standard 52.2-2017, filter efficiency is based on particle size removal efficiency (PSE).^11^ In other words, filter efficiency is the fraction of particles removed from air passing through the filter.^12^ Particles fall into three size ranges: E1 0.30-1.0 µm; E2 1.0-3.0 µm; and E3 3.0-10.0 µm. The PSEs in these size ranges are used to determine the MERV rating, which ranges from MERV 1 to MERV 16. Another common filter is a High Efficiency Particulate Air (HEPA) filter; the filter efficiency of a HEPA filter is better than MERV 16.^12^ Based on these dates, studies and recommendations may report a variety of filter efficiencies from EU, F, MERV, and HEPA.

In January 2021, ASHRAE^5^ made recommendations for reducing airborne infectious aerosol exposure, which included the use of MERV 13 or higher filters for air recirculated by HVAC systems. When accessed in April 2021, the ASHRAE website indicated that MERV 13 is recommended but MERV 14 or better is preferred.^13^ In March 2021, the WHO released a roadmap concerning indoor ventilation during COVID-19.^14^ Based on a scoping review that identified six studies specific to SARS-CoV-2 and review of technical guidance by leading international HVAC organizations, the roadmap presents standards for healthcare, non-residential and residential environments based on type of ventilation (natural vs. mechanical). In specific situations (e.g., depending on ventilation rate and airflow patterns, system designs with air recirculation modes or heat recovery), HEPA or MERV 14 filters are recommended.^14^

This systematic review considered whether virus transmission is affected by HVAC design features, particularly, filtration. In this review, an extensive and comprehensive search of the literature was conducted to identify and synthesize published research evaluating the effectiveness of filtration in reducing virus transmission. The insight drawn from this review could help answer questions of the utility of filtration to mitigate the transmission of SARS-CoV-2 in mechanically ventilated indoor environments. Further, understanding the association between filter efficiency and infection risk could inform control measures. Finally, a comprehensive synthesis of the existing scientific literature can identify gaps and guide priorities for future research.

## Methods

This systematic review addressed the research question: is virus transmission affected by HVAC design features, particularly, filtration? The current review was part of a larger research program to review literature on HVAC design features (ventilation, filtration, ultraviolet radiation, and humidity) and virus transmission. Results for other HVAC design features of interest are reported separately due to the volume and heterogeneity of the research. The systematic review is registered.^15^ An *a priori* protocol was developed and is publicly available.^16^ The standards for the conduct of systematic reviews defined by the international Cochrane organization^17^ were followed with modifications for questions of etiology.^18^ The review is reported according to accepted reporting standards.^19^

### Search Strategy

From inception to June 2020, three electronic databases (Ovid MEDLINE, Compendex, Web of Science Core) were searched by a research librarian (GMT) using concepts related to virus, transmission, and HVAC. The Ovid MEDLINE search strategy is detailed in Table 1. Prior to implementing the searches, the strategies were peer-reviewed by two librarians (TL, AH). In January 2021, the search was updated. Reference lists of all relevant papers were screened, in addition to relevant review articles. Conference abstracts were identified through Compendex and Web of Science and were not included, but literature was searched to see whether any potentially relevant abstracts had been published as complete papers. Limits by year or language of publication were not placed on the search; however, only English-language studies were included due to the volume of available literature and resource constraints. EndNote was used to manage references with duplicate records removed prior to screening.

**Table 1:**
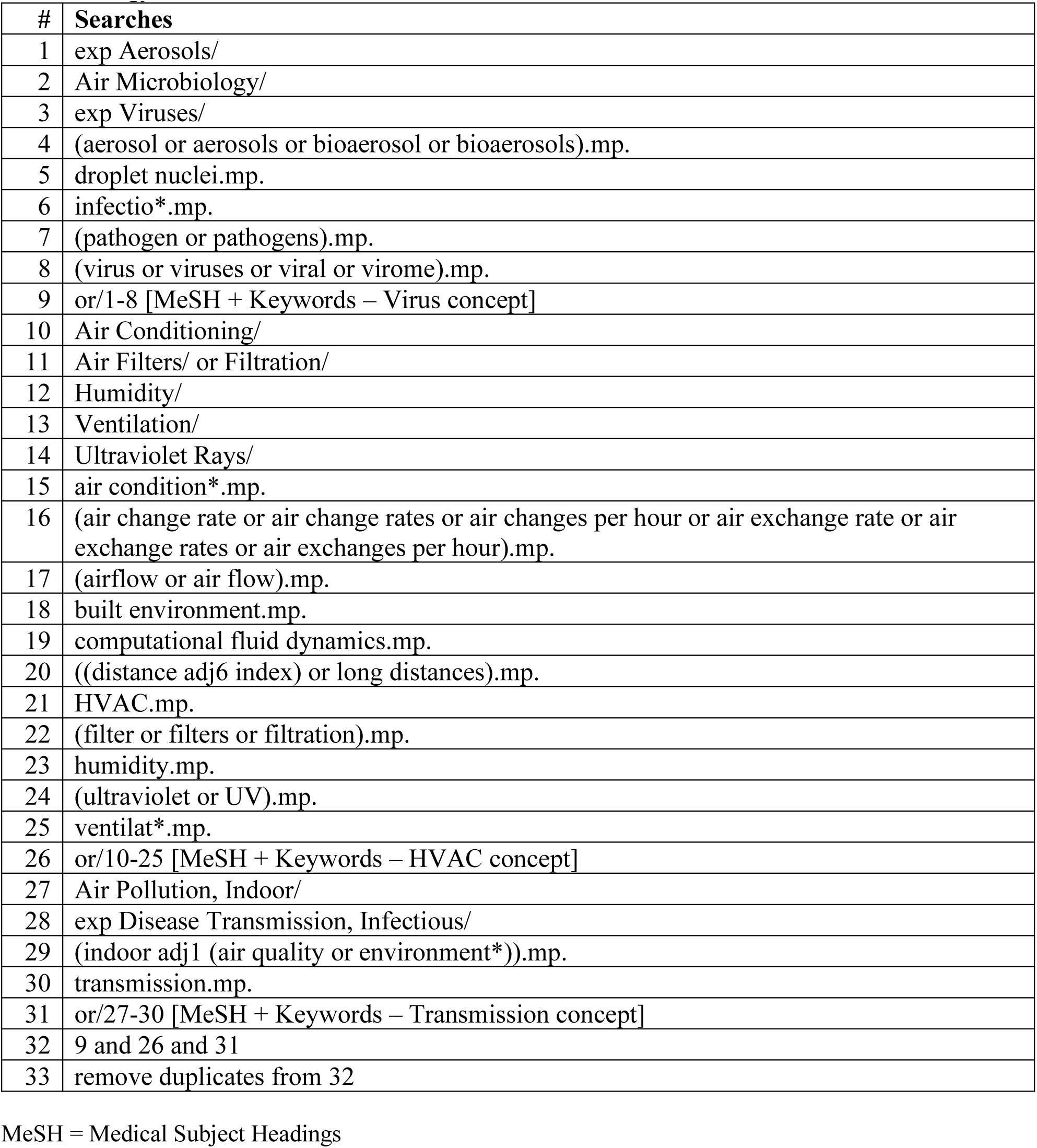
Search Strategy for Ovid MEDLINE^16^. Database: Ovid MEDLINE(R) ALL 1946 to Present Search Strategy:

| # | Searches |
| --- | --- |
| 1 | exp Aerosols/ |
| 2 | Air Microbiology/ |
| 3 | exp Viruses/ |
| 4 | (aerosol or aerosols or bioaerosol or bioaerosols).mp. |
| 5 | droplet nuclei.mp. |
| 6 | infectio*.mp. |
| 7 | (pathogen or pathogens).mp. |
| 8 | (virus or viruses or viral or virome).mp. |
| 9 | or/1-8 [MeSH + Keywords – Virus concept] |
| 10 | Air Conditioning/ |
| 11 | Air Filters/ or Filtration/ |
| 12 | Humidity/ |
| 13 | Ventilation/ |
| 14 | Ultraviolet Rays/ |
| 15 | air condition*.mp. |
| 16 | (air change rate or air change rates or air changes per hour or air exchange rate or air exchange rates or air exchanges per hour).mp. |
| 17 | (airflow or air flow).mp. |
| 18 | built environment.mp. |
| 19 | computational fluid dynamics.mp. |
| 20 | ((distance adj6 index) or long distances).mp. |
| 21 | HVAC.mp. |
| 22 | (filter or filters or filtration).mp. |
| 23 | humidity.mp. |
| 24 | (ultraviolet or UV).mp. |
| 25 | ventilat*.mp. |
| 26 | or/10-25 [MeSH + Keywords – HVAC concept] |
| 27 | Air Pollution, Indoor/ |
| 28 | exp Disease Transmission, Infectious/ |
| 29 | (indoor adj1 (air quality or environment*)).mp. |
| 30 | transmission.mp. |
| 31 | or/27-30 [MeSH + Keywords – Transmission concept] |
| 32 | 9 and 26 and 31 |
| 33 | remove duplicates from 32 |

MeSH = Medical Subject Headings

### Study Selection

Study selection had two stages. First, two reviewers independently screened titles and abstracts of references identified by electronic database searches. Relevance of each record was classified as Yes, Maybe, or No. Any conflicts between Yes/Maybe and No were resolved by one reviewer. To ensure consistency among the review team, pilot testing was conducted with three sets of studies (n=199 each). The review team met to discuss discrepancies and develop decision rules after each set of pilot screenings. Second, two reviewers independently reviewed full text articles and applied inclusion and exclusion criteria. Reviewers classified studies as Include or Exclude. Any conflicts between Include and Exclude were resolved by consensus of the review team. One reviewer resolved conflicts between different exclusion reasons. The second stage of screening was pilot tested with three sets of studies (n=30 each). The review team met to resolve discrepancies after each pilot round. The review team conducted screening using Covidence software.

### Inclusion and Exclusion Criteria

Inclusion and exclusion criteria are listed in Table 2. This systematic review was part of a larger effort to examine different HVAC design features and virus transmission. While all design features were included in the search and screening process at once, only studies evaluating filtration were synthesized here. A variety of agents were included in the search; however, studies of viruses or agents that simulated viruses were prioritized. Other agents (e.g., bacteria, fungi) would be included only if studies were not available that were specific to viruses. Studies using bacteriophages, which are viruses that infect bacterial cells,^20^ were included. This review is interested in studies of the indoor built environment (e.g., office, public, residential buildings) which had mechanical ventilation. Primary research that provided quantitative results of the correlation or association between filtration and virus transmission was included. No restrictions were placed on year of publication and only English-language, peer-reviewed publications were included.

**Table 2.** Inclusion and exclusion criteria for systematic review^16^.

| Item | Inclusion criteria | Exclusion criteria |
| --- | --- | --- |
| Agent | • Viruses<br>• Aerosols<br>• Bioaerosols<br>• Droplet nuclei<br>• Other pathogens (e.g., bacteria, fungi)<br>*We planned a staged process: if we identified studies specific to viruses for each HVAC design feature, we would not include other pathogens; however, for design features where we did not find studies specific to viruses, we would expand to other pathogens.* |  |
| HVAC | Design features relating to:<br>• Ventilation (ventilation rate, air changes per hour (ACH), air exchange, airflow pattern, pressurization)<br>• Filtration (air filtration, filter type, MERV rating, filter age and/or use, pressure drop, holding capacity, replacement, change frequency)<br>• Ultraviolet germicidal irradiation (UVGI; power, dose, uniformity of dose, flow rate, bioaerosol inactivation efficiency, location)<br>• Humidity or relative humidity | Examines HVAC / mechanical / or other ventilation mechanisms overall, but not by specific design features. |
| Setting | • Office buildings<br>• Public buildings (e.g., schools, day cares)<br>• Residential buildings<br>• Hospitals and other healthcare facilities (e.g., clinics)<br>• Transport vehicles (e.g., aircraft) or hubs (e.g., airports) | • Outdoor settings<br>• Indoor settings with natural ventilation |
| Outcomes | Quantitative data evaluating the correlation or association between virus transmission and above HVAC features | Qualitative data |
| Study design | Primary research, including:<br>• Epidemiological studies<br>• Observational studies (e.g., cohort, case-control, cross-sectional)<br>• Experimental studies (including human or animal)<br>• Modelling studies, including CFD | • Review articles<br>• Commentaries, opinion pieces<br>• Qualitative studies |
| Language | English<br>*We planned a staged process where we would include studies in languages other than English if we do not identify English language studies for specific HVAC design features or if we identified clusters of potentially relevant studies in another language.* |  |
| Year | No restrictions |  |
| Publication status | Published, peer-reviewed | Unpublished, not peer-reviewed |

CFD = computational fluid dynamics; HVAC = heating, ventilation, and air conditioning; MERV = minimum efficiency reporting value; UVGI = ultraviolet germicidal irradiation

### Risk of Bias Assessment

For experimental studies, risk of bias was assessed based on three key domains: selection bias, information bias and confounding.^21-22^ Reviewers assessed each domain as low, unclear, or high risk of bias using signalling questions^17^ from guidance documents for the different study types that were included; e.g., animal studies, laboratory experiments, epidemiological studies.^21-23^ For modelling studies, risk of bias was assessed using the following three key domains: definition, assumption, and validation.^23-24^ The domain definition considers model complexity and data sources, assumption considers the description and explanation of model assumptions, and validation considered model validation and sensitivity analysis.^24^ Reviewers assessed each domain as low, unclear, or high risk of bias based on signalling questions.^23-25^ Risk of bias items were pilot tested among three review authors. Then, two reviewers (GMT, BAF) applied the criteria independently to each relevant study and met to resolve discrepancies.

### Data Extraction

Reviewers extracted general information about the study (authors, year of publication, country of corresponding author, study design) and methods (setting, population [as applicable], agent studied, intervention set-up). Details on filtration parameters (where available) were extracted, including any filter efficiency ratings or classifications (such as MERV and EU ratings or HEPA) or filter efficiencies expressed as percentages, where applicable. More recent filter efficiencies are based on ASHRAE Standard 52.2-2017 and represent particle size removal efficiencies (PSE) with particles falling into three size ranges: E1 0.30-1.0 µm; E2 1.0-3.0 µm; and E3 3.0-10.0 µm.^11^ Efficiency of particle removal is usually represented by conventional filter efficiency ratings, such as MERV, EU, F, and HEPA. Quantitative data were extracted, as well as results of any tests of statistical significance related to filtration features. The primary outcome of interest was quantitative measures of the association between filtration and virus transmission. As such, data on actual transmission were extracted where available (i.e., infections), as well as information regarding viruses, filters, setting and experimental test set-up, and outcomes. The distinction between the efficiency of particle removal described above and the efficiency of virus removal is important. The efficiency of virus removal is calculated as the difference between the virus concentration upstream of the filter and the virus concentration downstream of the filter divided by the virus concentration upstream of the filter expressed as a percentage. Only one aerosolized virus study used terminology that referred explicitly to virus, viral filtration efficiency,^26^ while the remaining studies on virus removal used a variety of terms typical for particle removal: removal efficiency,^27^ filtrating efficiency,^28^ filter efficiency,^29^ and filter reduction efficiency.^30-31^ European ratings were presented with MERV rating conversions.^32^ Using a data extraction form spreadsheet to ensure comprehensive and consistent capture of data, one reviewer extracted data and a second reviewer verified data for accuracy and completeness. Any discrepancies were discussed by the review team.

### Data Synthesis

As anticipated, meta-analysis was not possible due to heterogeneity across studies in terms of study design, filtration features examined, outcomes assessed, and reporting of results. Evidence tables were developed to describe the studies and their results, as well as a narrative synthesis of the results. To allow for meaningful synthesis and comparison across studies, the studies are presented and discussed in three groups: animal studies (Table 3), aerosolized virus studies (Table 4), and modelling studies (Table 5).

**Table 3.**
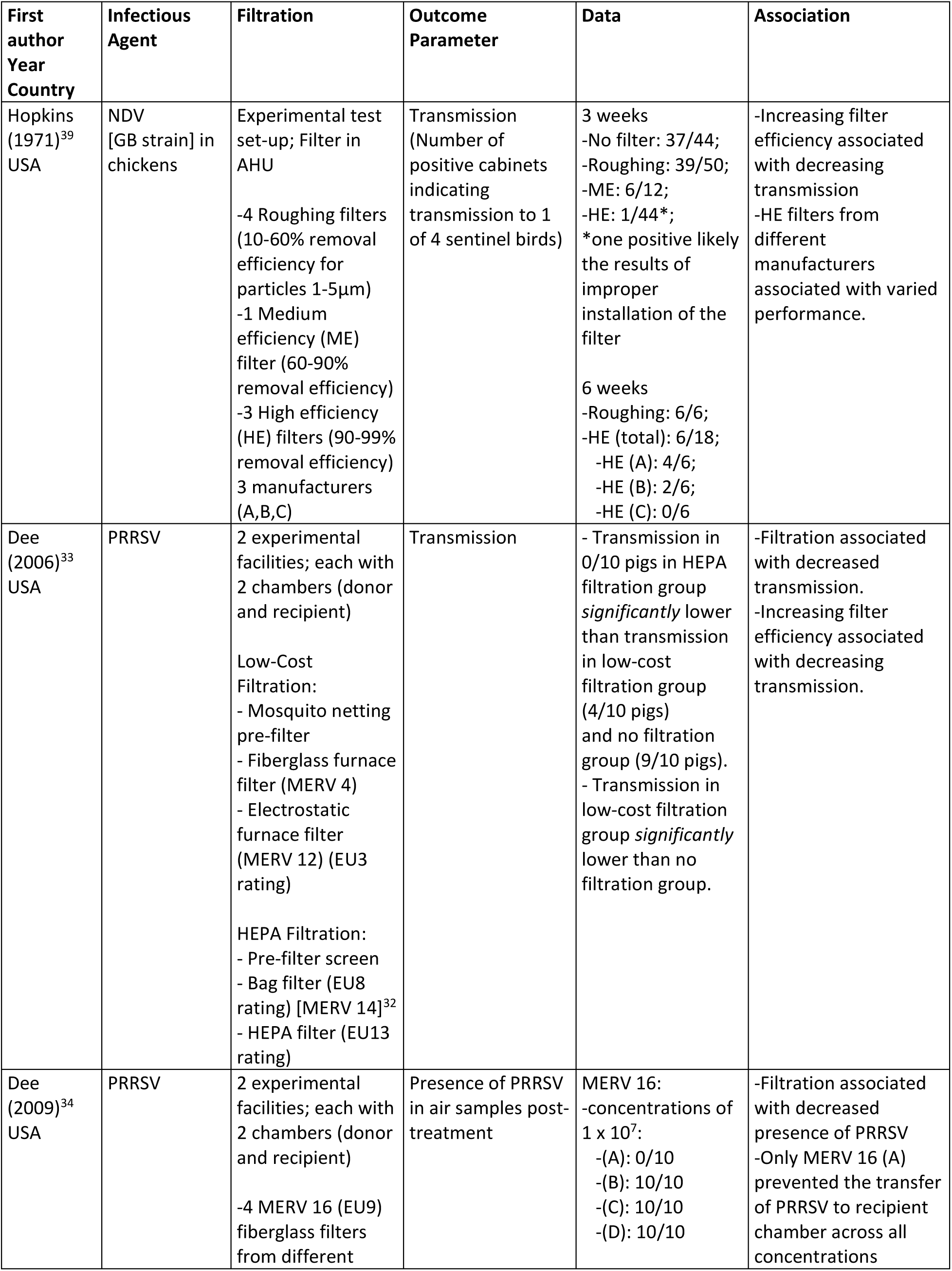
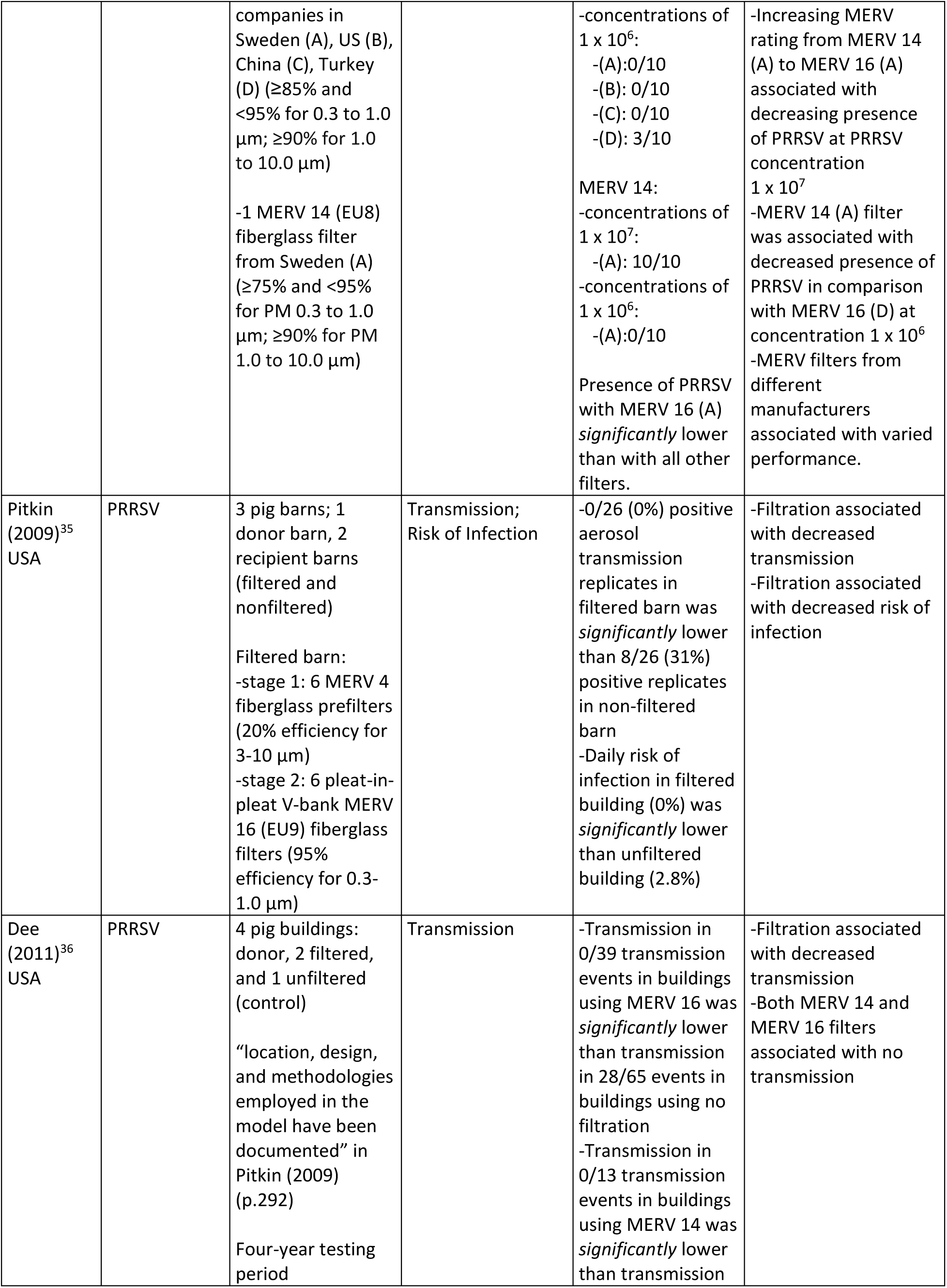
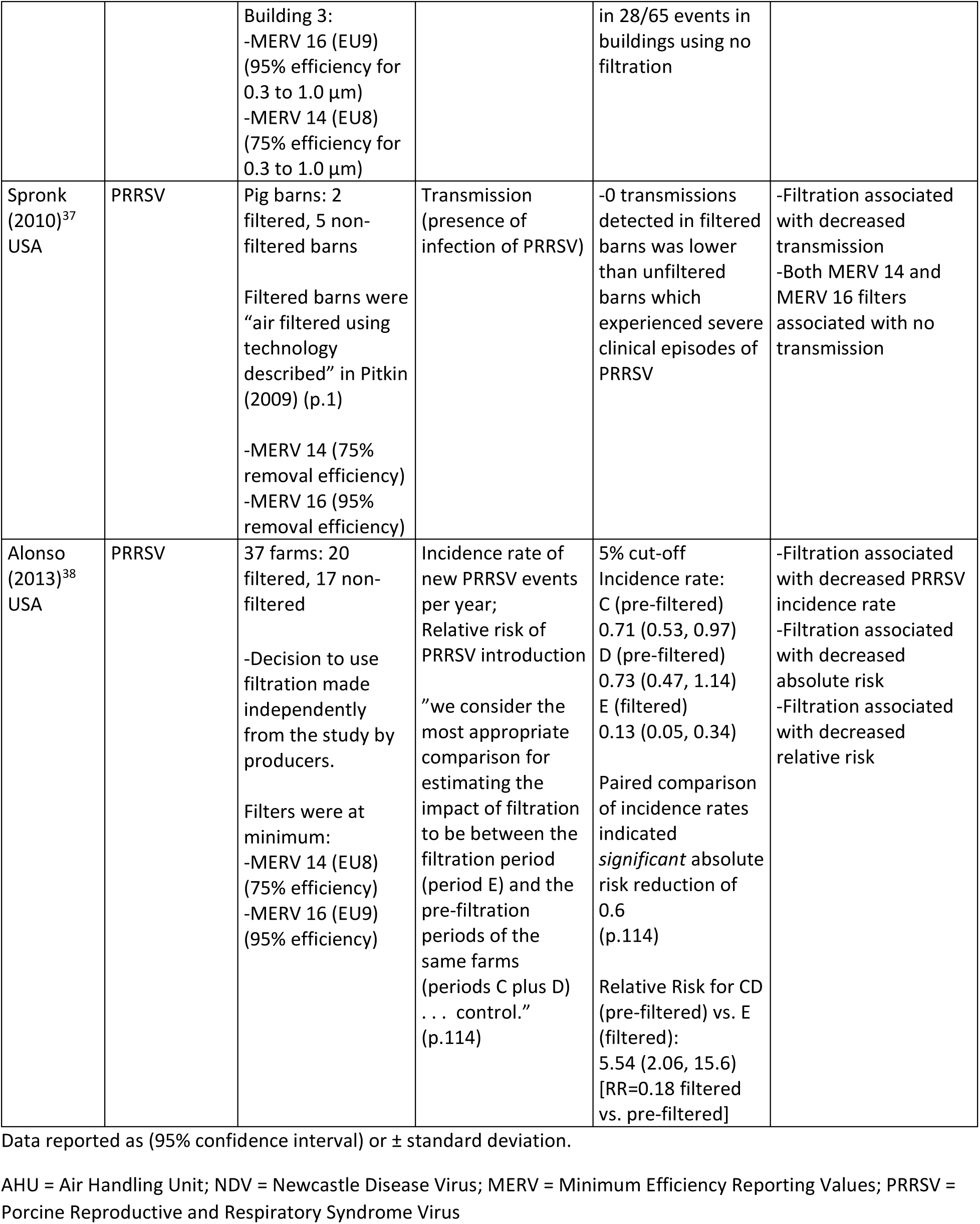
Animal Studies (n=7; listed in chronological order)

| First author<br>Year<br>Country | Infectious Agent | Filtration | Outcome Parameter | Data | Association |
| --- | --- | --- | --- | --- | --- |
| Hopkins (1971)<sup>39</sup><br>USA | NDV [GB strain] in chickens | Experimental test set-up; Filter in AHU<br>-4 Roughing filters (10-60% removal efficiency for particles 1-5µm)<br>-1 Medium efficiency (ME) filter (60-90% removal efficiency)<br>-3 High efficiency (HE) filters (90-99% removal efficiency)<br>3 manufacturers (A,B,C) | Transmission (Number of positive cabinets indicating transmission to 1 of 4 sentinel birds) | 3 weeks<br>-No filter: 37/44;<br>-Roughing: 39/50;<br>-ME: 6/12;<br>-HE: 1/44*;<br>*one positive likely the results of improper installation of the filter<br>6 weeks<br>-Roughing: 6/6;<br>-HE (total): 6/18;<br>-HE (A): 4/6;<br>-HE (B): 2/6;<br>-HE (C): 0/6 | -Increasing filter efficiency associated with decreasing transmission<br>-HE filters from different manufacturers associated with varied performance. |
| Dee (2006)<sup>33</sup><br>USA | PRRSV | 2 experimental facilities; each with 2 chambers (donor and recipient)<br>Low-Cost Filtration:<br>- Mosquito netting pre-filter<br>- Fiberglass furnace filter (MERV 4)<br>- Electrostatic furnace filter (MERV 12) (EU3 rating)<br>HEPA Filtration:<br>- Pre-filter screen<br>- Bag filter (EU8 rating) [MERV 14]<sup>32</sup><br>- HEPA filter (EU13 rating) | Transmission | - Transmission in 0/10 pigs in HEPA filtration group *significantly* lower than transmission in low-cost filtration group (4/10 pigs) and no filtration group (9/10 pigs).<br>- Transmission in low-cost filtration group *significantly* lower than no filtration group. | -Filtration associated with decreased transmission.<br>-Increasing filter efficiency associated with decreasing transmission. |
| Dee (2009)<sup>34</sup><br>USA | PRRSV | 2 experimental facilities; each with 2 chambers (donor and recipient)<br>-4 MERV 16 (EU9) fiberglass filters from different | Presence of PRRSV in air samples post-treatment | MERV 16:<br>-concentrations of $1 \times 10^7$:<br>-(A): 0/10<br>-(B): 10/10<br>-(C): 10/10<br>-(D): 10/10 | -Filtration associated with decreased presence of PRRSV<br>-Only MERV 16 (A) prevented the transfer of PRRSV to recipient chamber across all concentrations |
|  |  | companies in Sweden (A), US (B), China (C), Turkey (D) ($\geq 85\%$ and $< 95\%$ for 0.3 to 1.0 $\mu\text{m}$; $\geq 90\%$ for 1.0 to 10.0 $\mu\text{m}$)<br>-1 MERV 14 (EU8) fiberglass filter from Sweden (A) ($\geq 75\%$ and $< 95\%$ for PM 0.3 to 1.0 $\mu\text{m}$; $\geq 90\%$ for PM 1.0 to 10.0 $\mu\text{m}$) |  | -concentrations of $1 \times 10^6$:<br>-(A): 0/10<br>-(B): 0/10<br>-(C): 0/10<br>-(D): 3/10<br>MERV 14:<br>-concentrations of $1 \times 10^7$:<br>-(A): 10/10<br>-concentrations of $1 \times 10^6$:<br>-(A): 0/10<br>Presence of PRRSV with MERV 16 (A) *significantly* lower than with all other filters. | -Increasing MERV rating from MERV 14 (A) to MERV 16 (A) associated with decreasing presence of PRRSV at PRRSV concentration $1 \times 10^7$<br>-MERV 14 (A) filter was associated with decreased presence of PRRSV in comparison with MERV 16 (D) at concentration $1 \times 10^6$<br>-MERV filters from different manufacturers associated with varied performance. |
| Pitkin (2009)<sup>35</sup><br>USA | PRRSV | 3 pig barns; 1 donor barn, 2 recipient barns (filtered and nonfiltered)<br>Filtered barn:<br>-stage 1: 6 MERV 4 fiberglass prefilters (20% efficiency for 3-10 $\mu\text{m}$)<br>-stage 2: 6 pleat-in-pleat V-bank MERV 16 (EU9) fiberglass filters (95% efficiency for 0.3-1.0 $\mu\text{m}$) | Transmission;<br>Risk of Infection | -0/26 (0%) positive aerosol transmission replicates in filtered barn was *significantly* lower than 8/26 (31%) positive replicates in non-filtered barn<br>-Daily risk of infection in filtered building (0%) was *significantly* lower than unfiltered building (2.8%) | -Filtration associated with decreased transmission<br>-Filtration associated with decreased risk of infection |
| Dee (2011)<sup>36</sup><br>USA | PRRSV | 4 pig buildings: donor, 2 filtered, and 1 unfiltered (control)<br>"location, design, and methodologies employed in the model have been documented" in Pitkin (2009) (p.292)<br>Four-year testing period | Transmission | -Transmission in 0/39 transmission events in buildings using MERV 16 was *significantly* lower than transmission in 28/65 events in buildings using no filtration<br>-Transmission in 0/13 transmission events in buildings using MERV 14 was *significantly* lower than transmission | -Filtration associated with decreased transmission<br>-Both MERV 14 and MERV 16 filters associated with no transmission |
|  |  | Building 3:<br>-MERV 16 (EU9)<br>(95% efficiency for 0.3 to 1.0 µm)<br>-MERV 14 (EU8)<br>(75% efficiency for 0.3 to 1.0 µm) |  | in 28/65 events in buildings using no filtration |  |
| Spronk (2010)<sup>37</sup><br>USA | PRRSV | Pig barns: 2 filtered, 5 non-filtered barns<br>Filtered barns were “air filtered using technology described” in Pitkin (2009) (p.1)<br>-MERV 14 (75% removal efficiency)<br>-MERV 16 (95% removal efficiency) | Transmission (presence of infection of PRRSV) | -0 transmissions detected in filtered barns was lower than unfiltered barns which experienced severe clinical episodes of PRRSV | -Filtration associated with decreased transmission<br>-Both MERV 14 and MERV 16 filters associated with no transmission |
| Alonso (2013)<sup>38</sup><br>USA | PRRSV | 37 farms: 20 filtered, 17 non-filtered<br>-Decision to use filtration made independently from the study by producers.<br>Filters were at minimum:<br>-MERV 14 (EU8) (75% efficiency)<br>-MERV 16 (EU9) (95% efficiency) | Incidence rate of new PRRSV events per year;<br>Relative risk of PRRSV introduction<br>“we consider the most appropriate comparison for estimating the impact of filtration to be between the filtration period (period E) and the pre-filtration periods of the same farms (periods C plus D) . . . control.” (p.114) | 5% cut-off<br>Incidence rate:<br>C (pre-filtered) 0.71 (0.53, 0.97)<br>D (pre-filtered) 0.73 (0.47, 1.14)<br>E (filtered) 0.13 (0.05, 0.34)<br>Paired comparison of incidence rates indicated *significant* absolute risk reduction of 0.6 (p.114)<br>Relative Risk for CD (pre-filtered) vs. E (filtered): 5.54 (2.06, 15.6)<br>[RR=0.18 filtered vs. pre-filtered] | -Filtration associated with decreased PRRSV incidence rate<br>-Filtration associated with decreased absolute risk<br>-Filtration associated with decreased relative risk |

Data reported as (95% confidence interval) or ± standard deviation.
AHU = Air Handling Unit; NDV = Newcastle Disease Virus; MERV = Minimum Efficiency Reporting Values; PRRSV = Porcine Reproductive and Respiratory Syndrome Virus

**Table 4.**
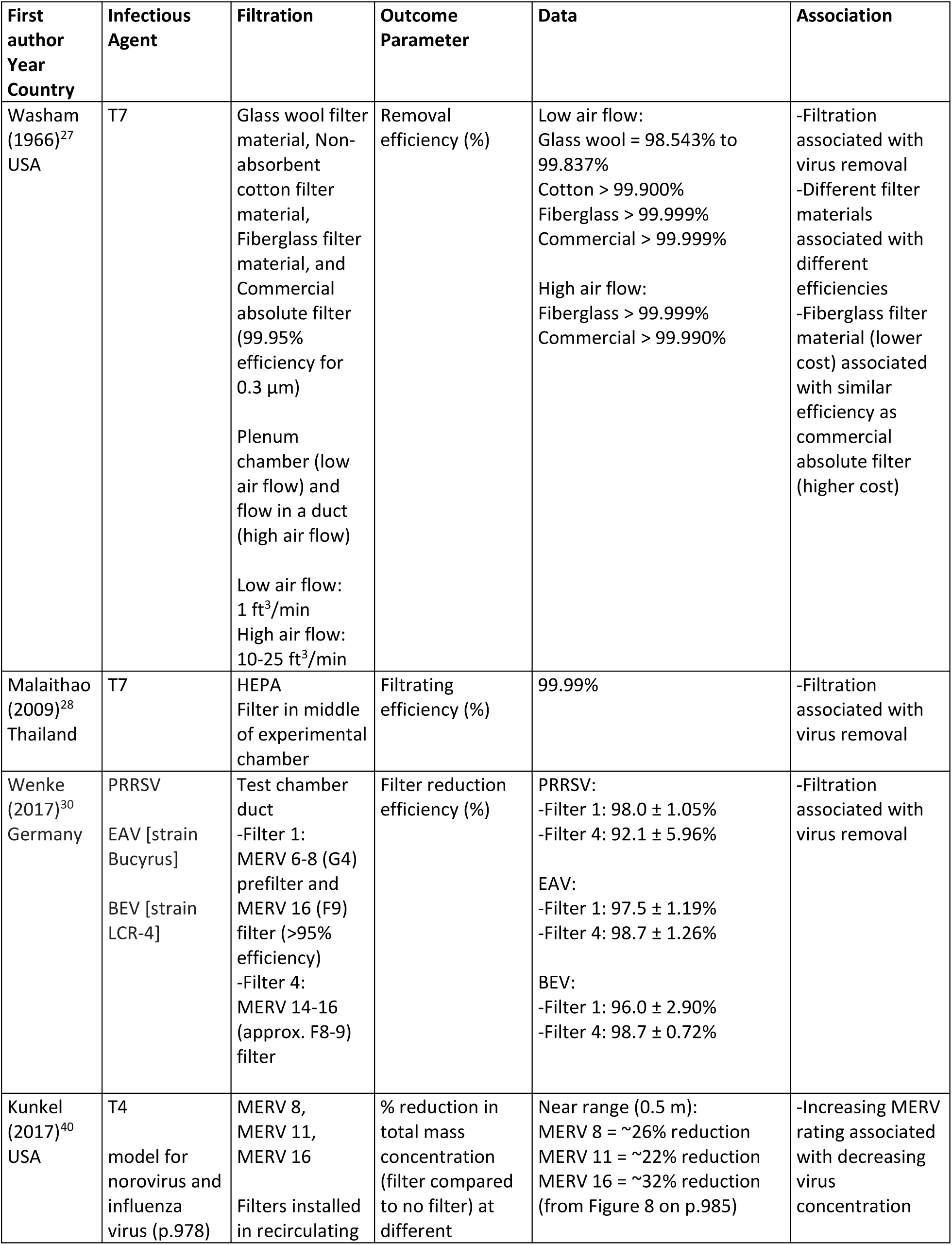
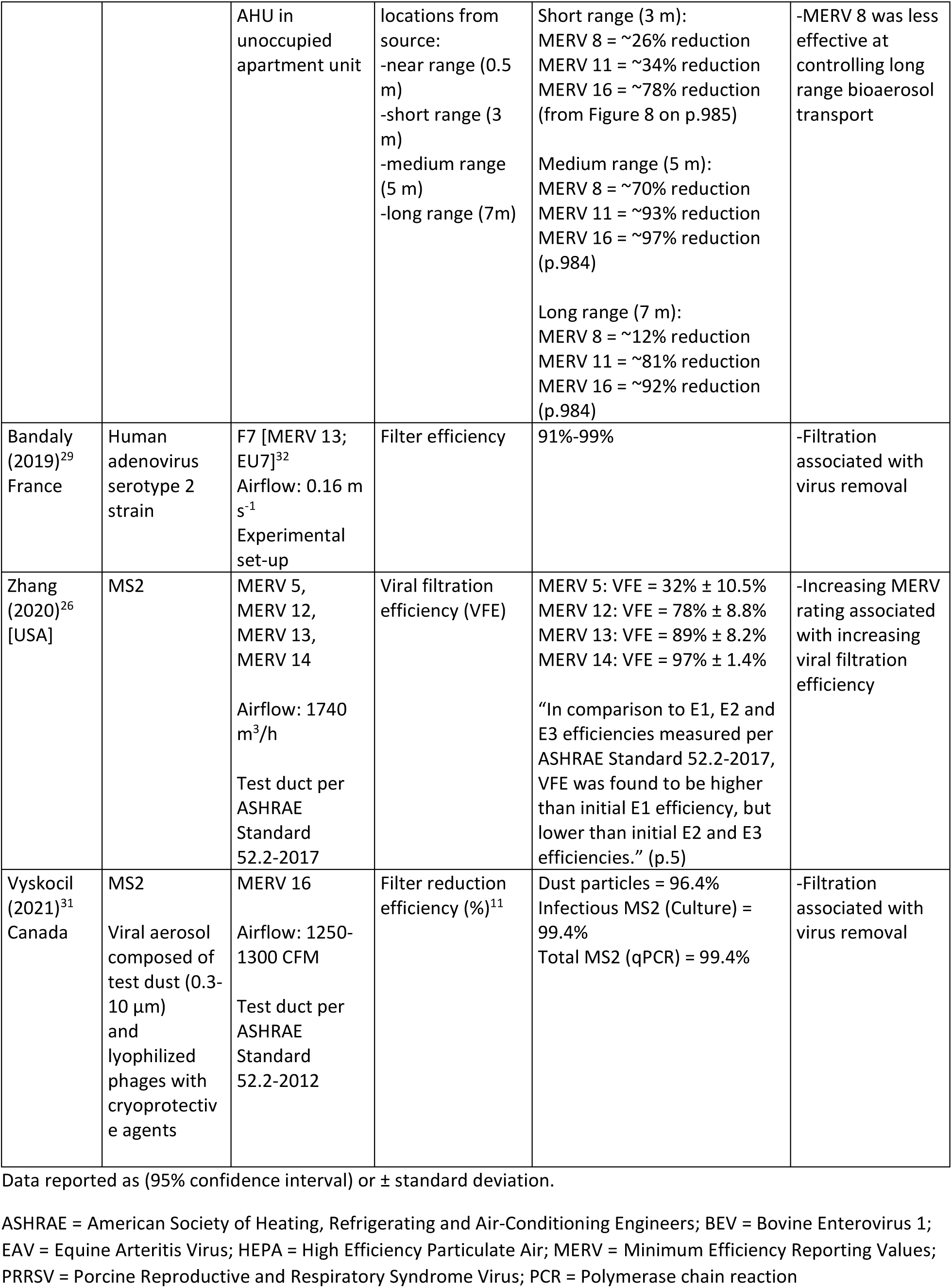
Aerosolized Virus Studies (n=7; listed in chronological order)

| First author<br>Year<br>Country | Infectious Agent | Filtration | Outcome Parameter | Data | Association |
| --- | --- | --- | --- | --- | --- |
| Washam (1966)<sup>27</sup><br>USA | T7 | Glass wool filter material, Non-absorbent cotton filter material, Fiberglass filter material, and Commercial absolute filter (99.95% efficiency for 0.3 µm)<br>Plenum chamber (low air flow) and flow in a duct (high air flow)<br>Low air flow: 1 ft<sup>3</sup>/min<br>High air flow: 10-25 ft<sup>3</sup>/min | Removal efficiency (%) | Low air flow:<br>Glass wool = 98.543% to 99.837%<br>Cotton > 99.900%<br>Fiberglass > 99.999%<br>Commercial > 99.999%<br>High air flow:<br>Fiberglass > 99.999%<br>Commercial > 99.990% | -Filtration associated with virus removal<br>-Different filter materials associated with different efficiencies<br>-Fiberglass filter material (lower cost) associated with similar efficiency as commercial absolute filter (higher cost) |
| Malaithao (2009)<sup>28</sup><br>Thailand | T7 | HEPA Filter in middle of experimental chamber | Filtrating efficiency (%) | 99.99% | -Filtration associated with virus removal |
| Wenke (2017)<sup>30</sup><br>Germany | PRRSV<br>EAV [strain Bucyrus]<br>BEV [strain LCR-4] | Test chamber duct<br>-Filter 1: MERV 6-8 (G4) prefilter and MERV 16 (F9) filter (>95% efficiency)<br>-Filter 4: MERV 14-16 (approx. F8-9) filter | Filter reduction efficiency (%) | PRRSV:<br>-Filter 1: 98.0 ± 1.05%<br>-Filter 4: 92.1 ± 5.96%<br>EAV:<br>-Filter 1: 97.5 ± 1.19%<br>-Filter 4: 98.7 ± 1.26%<br>BEV:<br>-Filter 1: 96.0 ± 2.90%<br>-Filter 4: 98.7 ± 0.72% | -Filtration associated with virus removal |
| Kunkel (2017)<sup>40</sup><br>USA | T4<br>model for norovirus and influenza virus (p.978) | MERV 8, MERV 11, MERV 16<br>Filters installed in recirculating | % reduction in total mass concentration (filter compared to no filter) at different | Near range (0.5 m):<br>MERV 8 = ~26% reduction<br>MERV 11 = ~22% reduction<br>MERV 16 = ~32% reduction (from Figure 8 on p.985) | -Increasing MERV rating associated with decreasing virus concentration |
|  |  | AHU in unoccupied apartment unit | locations from source:<br>-near range (0.5 m)<br>-short range (3 m)<br>-medium range (5 m)<br>-long range (7m) | Short range (3 m):<br>MERV 8 = ~26% reduction<br>MERV 11 = ~34% reduction<br>MERV 16 = ~78% reduction<br>(from Figure 8 on p.985)<br>Medium range (5 m):<br>MERV 8 = ~70% reduction<br>MERV 11 = ~93% reduction<br>MERV 16 = ~97% reduction<br>(p.984)<br>Long range (7 m):<br>MERV 8 = ~12% reduction<br>MERV 11 = ~81% reduction<br>MERV 16 = ~92% reduction<br>(p.984) | -MERV 8 was less effective at controlling long range bioaerosol transport |
| Bandaly (2019)<sup>29</sup><br>France | Human adenovirus serotype 2 strain | F7 [MERV 13; EU7]<sup>32</sup><br>Airflow: 0.16 m s<sup>-1</sup><br>Experimental set-up | Filter efficiency | 91%-99% | -Filtration associated with virus removal |
| Zhang (2020)<sup>26</sup><br>[USA] | MS2 | MERV 5, MERV 12, MERV 13, MERV 14<br>Airflow: 1740 m<sup>3</sup>/h<br>Test duct per ASHRAE Standard 52.2-2017 | Viral filtration efficiency (VFE) | MERV 5: VFE = 32% ± 10.5%<br>MERV 12: VFE = 78% ± 8.8%<br>MERV 13: VFE = 89% ± 8.2%<br>MERV 14: VFE = 97% ± 1.4%<br>“In comparison to E1, E2 and E3 efficiencies measured per ASHRAE Standard 52.2-2017, VFE was found to be higher than initial E1 efficiency, but lower than initial E2 and E3 efficiencies.” (p.5) | -Increasing MERV rating associated with increasing viral filtration efficiency |
| Vyskocil (2021)<sup>31</sup><br>Canada | MS2<br>Viral aerosol composed of test dust (0.3-10 µm) and lyophilized phages with cryoprotective agents | MERV 16<br>Airflow: 1250-1300 CFM<br>Test duct per ASHRAE Standard 52.2-2012 | Filter reduction efficiency (%)<sup>11</sup> | Dust particles = 96.4%<br>Infectious MS2 (Culture) = 99.4%<br>Total MS2 (qPCR) = 99.4% | -Filtration associated with virus removal |

Data reported as (95% confidence interval) or ± standard deviation.
ASHRAE = American Society of Heating, Refrigerating and Air-Conditioning Engineers; BEV = Bovine Enterovirus 1; EAV = Equine Arteritis Virus; HEPA = High Efficiency Particulate Air; MERV = Minimum Efficiency Reporting Values; PRRSV = Porcine Reproductive and Respiratory Syndrome Virus; PCR = Polymerase chain reaction

**Table 5.**
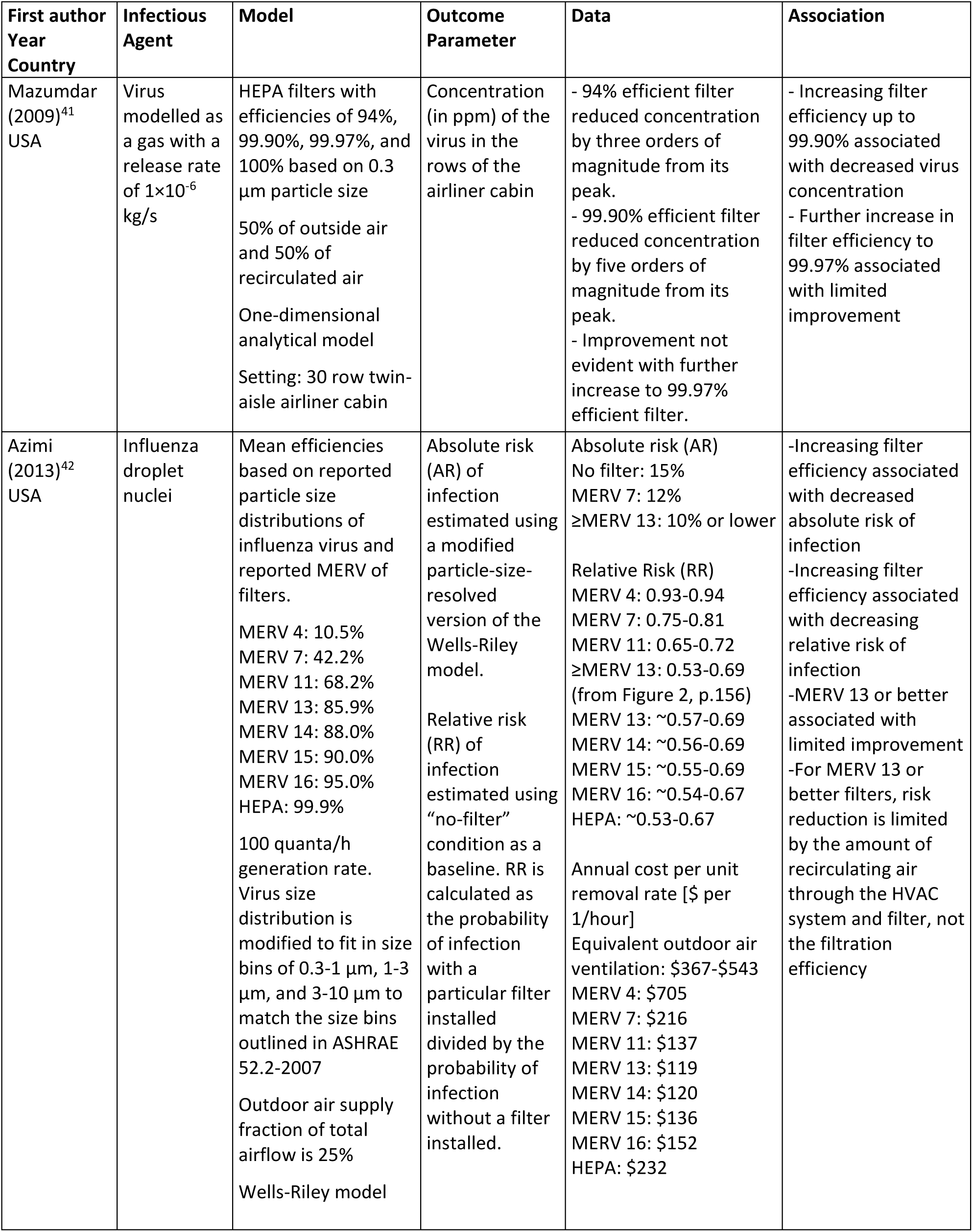
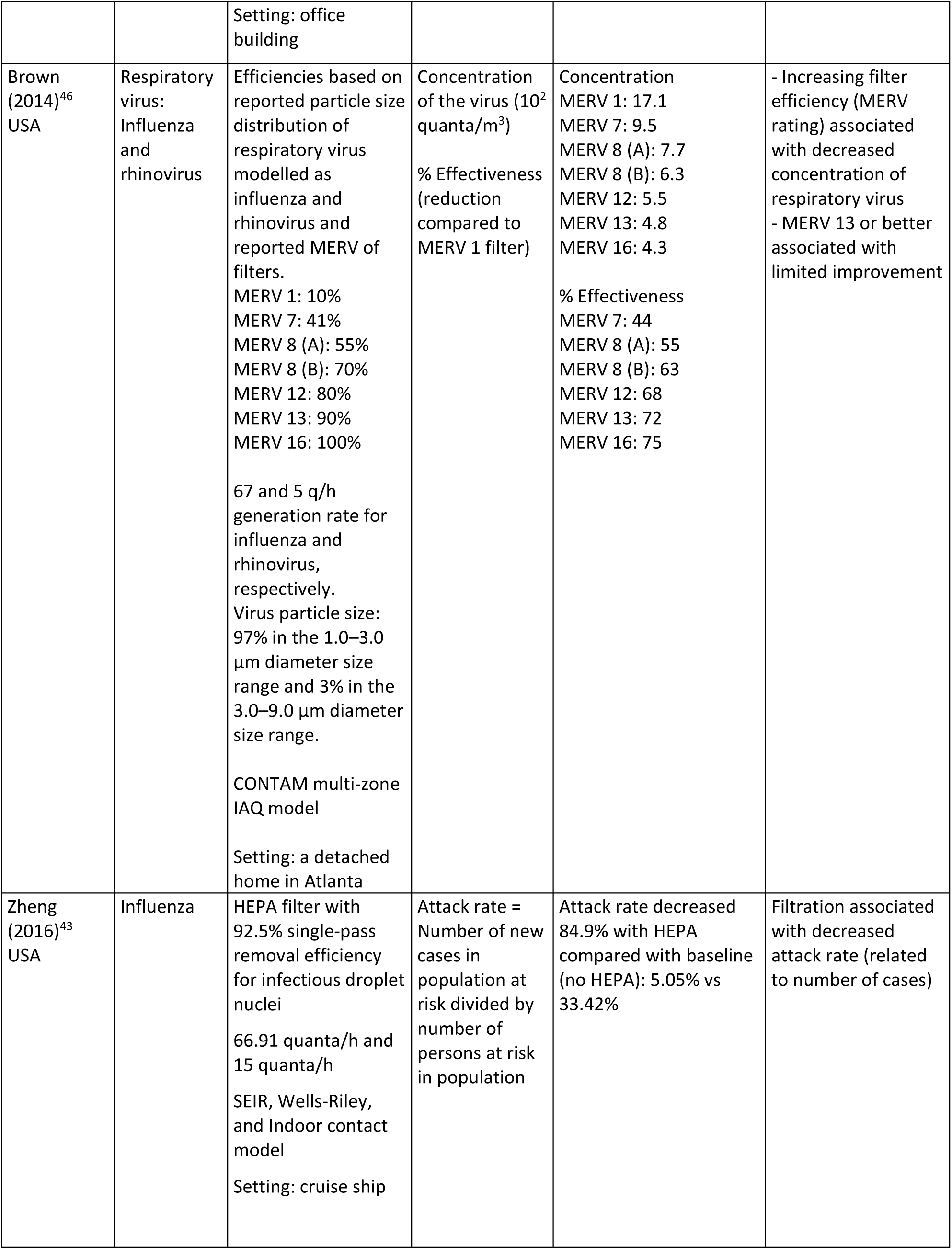
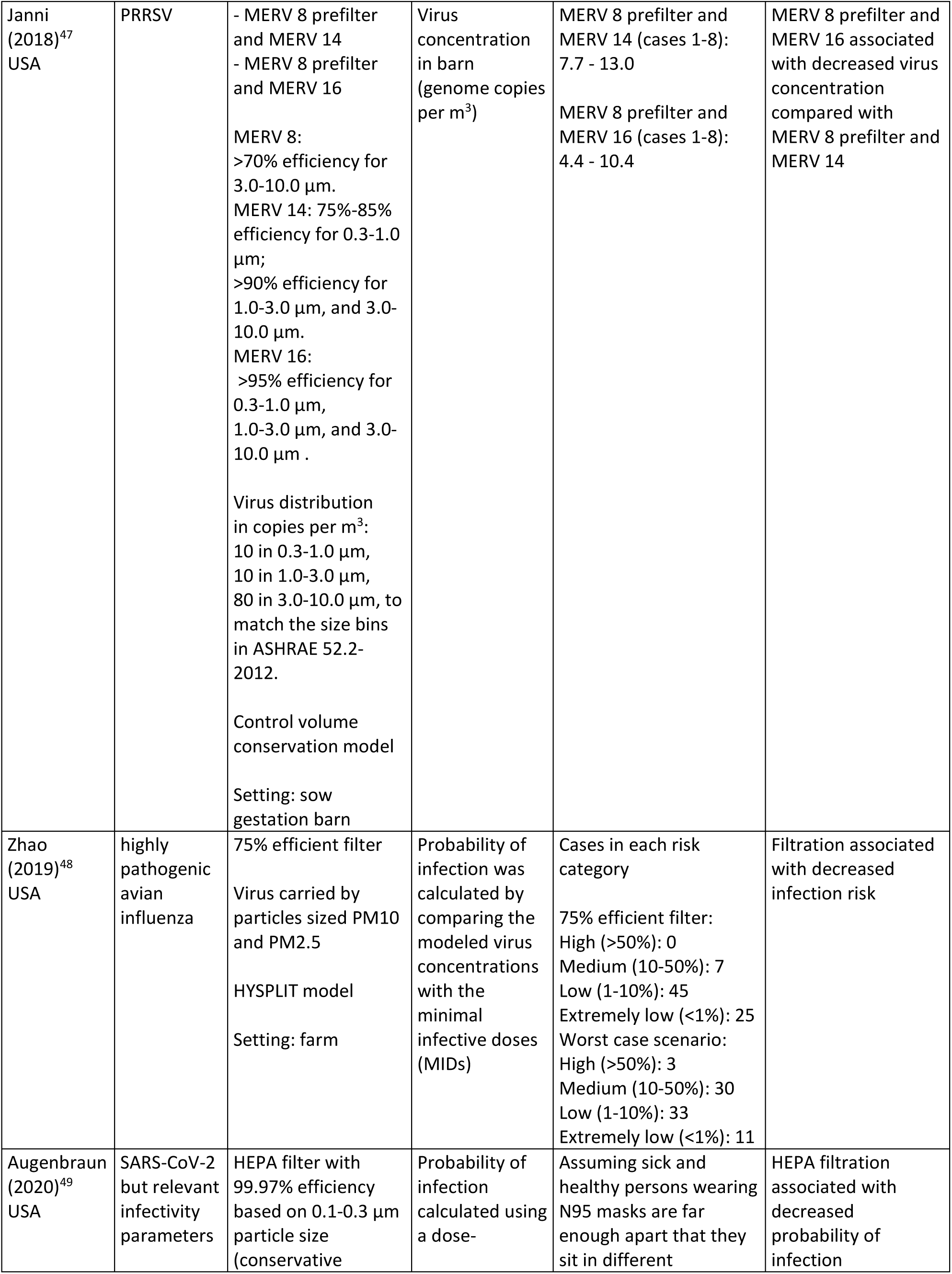
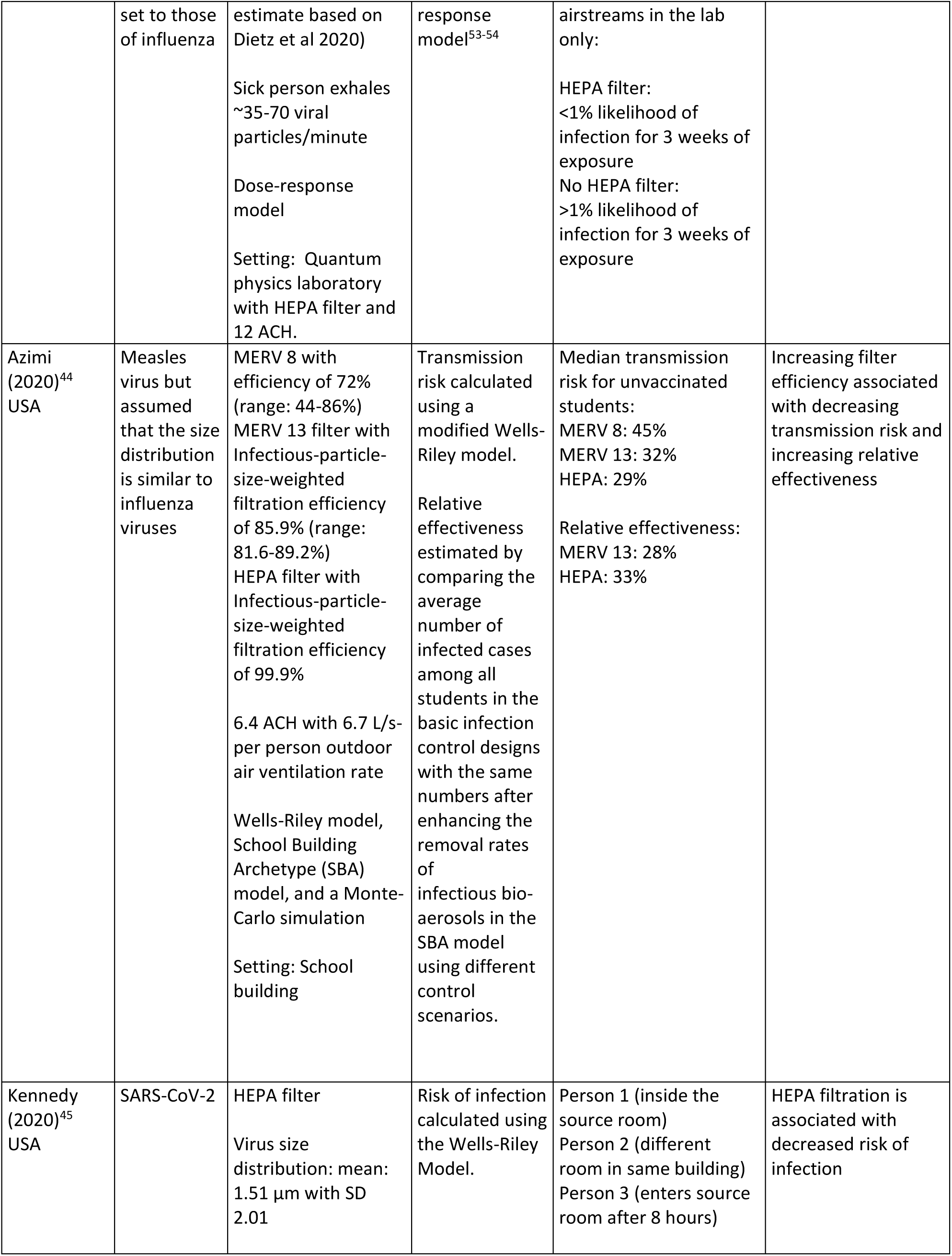
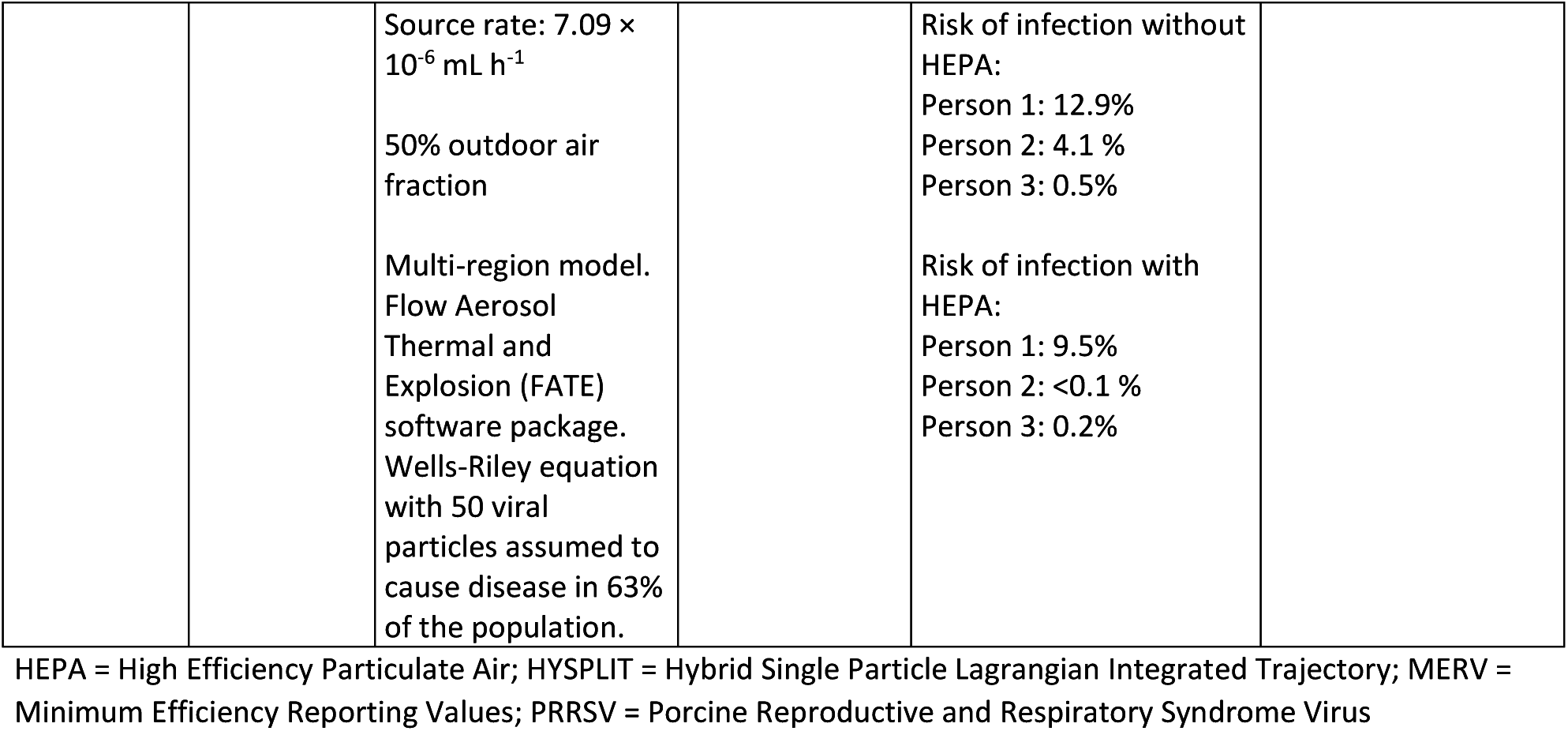
Modelling Studies (n=9; listed in chronological order)

| First author<br>Year<br>Country | Infectious<br>Agent | Model | Outcome<br>Parameter | Data | Association |
| --- | --- | --- | --- | --- | --- |
| Mazumdar<br>(2009)<sup>41</sup><br>USA | Virus<br>modelled as<br>a gas with a<br>release rate<br>of $1 \times 10^{-6}$<br>kg/s | HEPA filters with<br>efficiencies of 94%,<br>99.90%, 99.97%, and<br>100% based on 0.3<br>$\mu\text{m}$ particle size<br>50% of outside air<br>and 50% of<br>recirculated air<br>One-dimensional<br>analytical model<br>Setting: 30 row twin-<br>aisle airliner cabin | Concentration<br>(in ppm) of the<br>virus in the<br>rows of the<br>airliner cabin | - 94% efficient filter<br>reduced concentration<br>by three orders of<br>magnitude from its<br>peak.<br>- 99.90% efficient filter<br>reduced concentration<br>by five orders of<br>magnitude from its<br>peak.<br>- Improvement not<br>evident with further<br>increase to 99.97%<br>efficient filter. | - Increasing filter<br>efficiency up to<br>99.90% associated<br>with decreased virus<br>concentration<br>- Further increase in<br>filter efficiency to<br>99.97% associated<br>with limited<br>improvement |
| Azimi<br>(2013)<sup>42</sup><br>USA | Influenza<br>droplet<br>nuclei | Mean efficiencies<br>based on reported<br>particle size<br>distributions of<br>influenza virus and<br>reported MERV of<br>filters.<br>MERV 4: 10.5%<br>MERV 7: 42.2%<br>MERV 11: 68.2%<br>MERV 13: 85.9%<br>MERV 14: 88.0%<br>MERV 15: 90.0%<br>MERV 16: 95.0%<br>HEPA: 99.9%<br>100 quanta/h<br>generation rate.<br>Virus size<br>distribution is<br>modified to fit in size<br>bins of 0.3-1 $\mu\text{m}$, 1-3<br>$\mu\text{m}$, and 3-10 $\mu\text{m}$ to<br>match the size bins<br>outlined in ASHRAE<br>52.2-2007<br>Outdoor air supply<br>fraction of total<br>airflow is 25%<br>Wells-Riley model | Absolute risk<br>(AR) of<br>infection<br>estimated using<br>a modified<br>particle-size-<br>resolved<br>version of the<br>Wells-Riley<br>model.<br>Relative risk<br>(RR) of<br>infection<br>estimated using<br>“no-filter”<br>condition as a<br>baseline. RR is<br>calculated as<br>the probability<br>of infection<br>with a<br>particular filter<br>installed<br>divided by the<br>probability of<br>infection<br>without a filter<br>installed. | Absolute risk (AR)<br>No filter: 15%<br>MERV 7: 12%<br>≥MERV 13: 10% or lower<br>Relative Risk (RR)<br>MERV 4: 0.93-0.94<br>MERV 7: 0.75-0.81<br>MERV 11: 0.65-0.72<br>≥MERV 13: 0.53-0.69<br>(from Figure 2, p.156)<br>MERV 13: ∼0.57-0.69<br>MERV 14: ∼0.56-0.69<br>MERV 15: ∼0.55-0.69<br>MERV 16: ∼0.54-0.67<br>HEPA: ∼0.53-0.67<br>Annual cost per unit<br>removal rate [\$ per<br>1/hour]<br>Equivalent outdoor air<br>ventilation: \$367-\$543<br>MERV 4: \$705<br>MERV 7: \$216<br>MERV 11: \$137<br>MERV 13: \$119<br>MERV 14: \$120<br>MERV 15: \$136<br>MERV 16: \$152<br>HEPA: \$232 | -Increasing filter<br>efficiency associated<br>with decreased<br>absolute risk of<br>infection<br>-Increasing filter<br>efficiency associated<br>with decreasing<br>relative risk of<br>infection<br>-MERV 13 or better<br>associated with<br>limited improvement<br>-For MERV 13 or<br>better filters, risk<br>reduction is limited<br>by the amount of<br>recirculating air<br>through the HVAC<br>system and filter, not<br>the filtration<br>efficiency |
|  |  | Setting: office building |  |  |  |
| Brown (2014)<sup>46</sup><br>USA | Respiratory virus: Influenza and rhinovirus | Efficiencies based on reported particle size distribution of respiratory virus modelled as influenza and rhinovirus and reported MERV of filters.<br>MERV 1: 10%<br>MERV 7: 41%<br>MERV 8 (A): 55%<br>MERV 8 (B): 70%<br>MERV 12: 80%<br>MERV 13: 90%<br>MERV 16: 100%<br>67 and 5 q/h generation rate for influenza and rhinovirus, respectively.<br>Virus particle size: 97% in the 1.0–3.0 µm diameter size range and 3% in the 3.0–9.0 µm diameter size range.<br>CONTAM multi-zone IAQ model<br>Setting: a detached home in Atlanta | Concentration of the virus (10<sup>2</sup> quanta/m<sup>3</sup>)<br>% Effectiveness (reduction compared to MERV 1 filter) | Concentration<br>MERV 1: 17.1<br>MERV 7: 9.5<br>MERV 8 (A): 7.7<br>MERV 8 (B): 6.3<br>MERV 12: 5.5<br>MERV 13: 4.8<br>MERV 16: 4.3<br>% Effectiveness<br>MERV 7: 44<br>MERV 8 (A): 55<br>MERV 8 (B): 63<br>MERV 12: 68<br>MERV 13: 72<br>MERV 16: 75 | - Increasing filter efficiency (MERV rating) associated with decreased concentration of respiratory virus<br>- MERV 13 or better associated with limited improvement |
| Zheng (2016)<sup>43</sup><br>USA | Influenza | HEPA filter with 92.5% single-pass removal efficiency for infectious droplet nuclei<br>66.91 quanta/h and 15 quanta/h<br>SEIR, Wells-Riley, and Indoor contact model<br>Setting: cruise ship | Attack rate = Number of new cases in population at risk divided by number of persons at risk in population | Attack rate decreased 84.9% with HEPA compared with baseline (no HEPA): 5.05% vs 33.42% | Filtration associated with decreased attack rate (related to number of cases) |
| Janni (2018)<sup>47</sup><br>USA | PRRSV | - MERV 8 prefilter and MERV 14<br>- MERV 8 prefilter and MERV 16<br>MERV 8:<br>>70% efficiency for 3.0-10.0 $\mu\text{m}$.<br>MERV 14: 75%-85% efficiency for 0.3-1.0 $\mu\text{m}$;<br>>90% efficiency for 1.0-3.0 $\mu\text{m}$, and 3.0-10.0 $\mu\text{m}$.<br>MERV 16:<br>>95% efficiency for 0.3-1.0 $\mu\text{m}$, 1.0-3.0 $\mu\text{m}$, and 3.0-10.0 $\mu\text{m}$.<br>Virus distribution in copies per $\text{m}^3$:<br>10 in 0.3-1.0 $\mu\text{m}$,<br>10 in 1.0-3.0 $\mu\text{m}$,<br>80 in 3.0-10.0 $\mu\text{m}$, to match the size bins in ASHRAE 52.2-2012.<br>Control volume conservation model<br>Setting: sow gestation barn | Virus concentration in barn (genome copies per $\text{m}^3$) | MERV 8 prefilter and MERV 14 (cases 1-8): 7.7 - 13.0<br>MERV 8 prefilter and MERV 16 (cases 1-8): 4.4 - 10.4 | MERV 8 prefilter and MERV 16 associated with decreased virus concentration compared with MERV 8 prefilter and MERV 14 |
| Zhao (2019)<sup>48</sup><br>USA | highly pathogenic avian influenza | 75% efficient filter<br>Virus carried by particles sized PM10 and PM2.5<br>HYSPLIT model<br>Setting: farm | Probability of infection was calculated by comparing the modeled virus concentrations with the minimal infective doses (MIDs) | Cases in each risk category<br>75% efficient filter:<br>High (>50%): 0<br>Medium (10-50%): 7<br>Low (1-10%): 45<br>Extremely low (<1%): 25<br>Worst case scenario:<br>High (>50%): 3<br>Medium (10-50%): 30<br>Low (1-10%): 33<br>Extremely low (<1%): 11 | Filtration associated with decreased infection risk |
| Augenbraun (2020)<sup>49</sup><br>USA | SARS-CoV-2 but relevant infectivity parameters | HEPA filter with 99.97% efficiency based on 0.1-0.3 $\mu\text{m}$ particle size (conservative | Probability of infection calculated using a dose- | Assuming sick and healthy persons wearing N95 masks are far enough apart that they sit in different | HEPA filtration associated with decreased probability of infection |
|  | set to those of influenza | estimate based on Dietz et al 2020)<br>Sick person exhales ~35-70 viral particles/minute<br>Dose-response model<br>Setting: Quantum physics laboratory with HEPA filter and 12 ACH. | response model<sup>53-54</sup> | airstreams in the lab only:<br>HEPA filter: <1% likelihood of infection for 3 weeks of exposure<br>No HEPA filter: >1% likelihood of infection for 3 weeks of exposure |  |
| Azimi (2020)<sup>44</sup><br>USA | Measles virus but assumed that the size distribution is similar to influenza viruses | MERV 8 with efficiency of 72% (range: 44-86%)<br>MERV 13 filter with Infectious-particle-size-weighted filtration efficiency of 85.9% (range: 81.6-89.2%)<br>HEPA filter with Infectious-particle-size-weighted filtration efficiency of 99.9%<br>6.4 ACH with 6.7 L/s-per person outdoor air ventilation rate<br>Wells-Riley model, School Building Archetype (SBA) model, and a Monte-Carlo simulation<br>Setting: School building | Transmission risk calculated using a modified Wells-Riley model.<br>Relative effectiveness estimated by comparing the average number of infected cases among all students in the basic infection control designs with the same numbers after enhancing the removal rates of infectious bio-aerosols in the SBA model using different control scenarios. | Median transmission risk for unvaccinated students:<br>MERV 8: 45%<br>MERV 13: 32%<br>HEPA: 29%<br>Relative effectiveness:<br>MERV 13: 28%<br>HEPA: 33% | Increasing filter efficiency associated with decreasing transmission risk and increasing relative effectiveness |
| Kennedy (2020)<sup>45</sup><br>USA | SARS-CoV-2 | HEPA filter<br>Virus size distribution: mean: 1.51 $\mu\text{m}$ with SD 2.01 | Risk of infection calculated using the Wells-Riley Model. | Person 1 (inside the source room)<br>Person 2 (different room in same building)<br>Person 3 (enters source room after 8 hours) | HEPA filtration is associated with decreased risk of infection |

|  |  |  |  |  |
| --- | --- | --- | --- | --- |
|  |  | Source rate: $7.09 \times 10^{-6} \text{ mL h}^{-1}$<br>50% outdoor air fraction<br>Multi-region model. Flow Aerosol Thermal and Explosion (FATE) software package. Wells-Riley equation with 50 viral particles assumed to cause disease in 63% of the population. |  | Risk of infection without HEPA:<br>Person 1: 12.9%<br>Person 2: 4.1 %<br>Person 3: 0.5%<br>Risk of infection with HEPA:<br>Person 1: 9.5%<br>Person 2: <0.1 %<br>Person 3: 0.2% |

HEPA = High Efficiency Particulate Air; HYSPLIT = Hybrid Single Particle Lagrangian Integrated Trajectory; MERV = Minimum Efficiency Reporting Values; PRRSV = Porcine Reproductive and Respiratory Syndrome Virus

## Results

12,177 unique citations were identified following searches executed using the search protocol; 2,428 were identified as potentially relevant following title/abstract screening and 568 met the inclusion criteria following full-text review (Figure 1). Fifty-eight of the 568 were relevant to filtration; 23 studies met the inclusion criteria for filtration and viruses following full-text review (Figure 1). Studies were published between 1966 and 2021 (median year 2014). The majority (n=19) of studies were conducted in the United States. Funding sources for the studies included national research funding organizations and initiatives (n=8), industry (n=4), foundations (n=2), university grants (n=1), filtration associations (n=1), and research networks (n=1). Six studies reported no funding or did not list funding sources.

**Figure 1.**
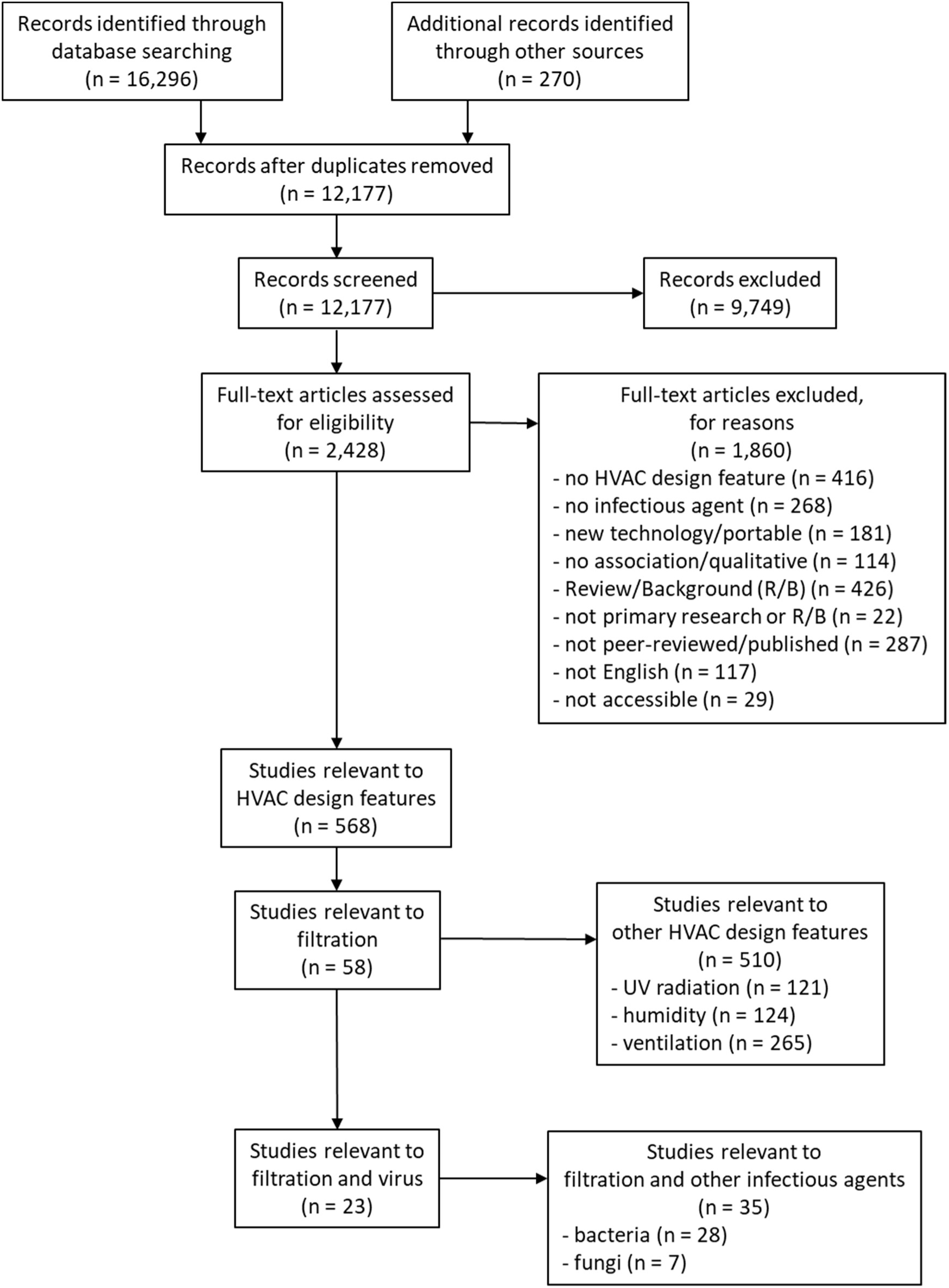
Flow of studies through the selection process (note: search was conducted for all HVAC design features but only studies of filtration are included in this manuscript)

In this review, studies were examined in three categories: animal studies (n=7), aerosolized virus studies (n=7), and modelling studies (n=9). Within the 23 studies, seven viruses and three bacteriophages were studied, including SARS-CoV-2 (COVID-19) (n=2), porcine reproductive and respiratory syndrome virus (PRRSV) (n=8), influenza (n=3), measles (n=1), Newcastle Disease Virus (NDV) (n=1), adenovirus (n=1), and avian influenza (n=1). Details of individual studies are provided in Tables 3, 4, and 5.

### Animal Studies (n=7)

Six studies evaluated the influence of filtration on PRRSV in pigs^33-38^ and one study examined NDV in chickens.^39^ Of the seven studies, three were performed using experimental test set-ups ups^33-34,39^ and four were performed in the field (i.e., pig barns).^35-38^

Five of the seven studies found that filtration was associated with decreased virus transmission (Table 3).^33,35-37,39^ Pitkin et al^35^ also found that filtration was associated with decreased daily risk of infections for PRRSV. One study found the use of filtration was associated with decreased presence of PRRSV downstream of the filter^34^ and another^38^ found the use of filtration was associated with decreased relative risk of PRRSV introductions.

Dee et al^33^ and Hopkins and Drury^39^ found that increased filter efficiency was associated with decreasing transmission. Hopkins and Drury^39^ documented that improper installation of a high efficiency filter allowed transmission to happen. Additionally, Dee et al^36^ found no difference in the transmission reduction between MERV 14 and MERV 16 with no transmission events in both situations. As well, two studies found that filter performance varied by manufacturer when testing similarly rated filters from different companies.^34,39^

### Aerosolized Virus Studies (n=7)

Five of seven aerosolized virus studies found that filtration was associated with virus removal using PRRSV,^30^ equine arteritis virus (EAV),^30^ bovine enterovirus (BEV),^30^ human adenovirus,^29^ and the bacteriophages T7^27-28^ and MS2.^31^ One study found that increasing MERV rating was associated with decreasing virus concentration for bacteriophage T4^40^ at different locations within an apartment unit, and another found that increasing MERV rating was associated with increasing viral filtration efficiency using bacteriophage MS2^26^ in a test duct set-up (Table 4).

Like the animal studies conducted by Dee et al^33^ and Hopkins and Drury,^39^ two aerosolized studies found increasing MERV ratings to be associated with increased virus removal efficiencies.^26,40^ Zhang et al^26^ noted that the filter efficiency associated with E1 (i.e., 0.30-1.0 µm; the smallest particle size range for MERV classification) was a good approximation of the viral filtration efficiency. In addition, Washam et al,^27^ prior to the use of MERV ratings as developed in 1999, identified cost differences between filters of similar efficiency.

### Modelling Studies (n=9)

Nine of the filtration studies were modelling, employing the following models: one-dimensional analytical model,^41^ Wells-Riley model,^42-45^ CONTAM multi-zone IAQ model,^46^ Susceptible-Exposed-Infectious-Recovered (SEIR) model,^43^ an indoor contact model,^43^ a control volume conservation model,^47^ Hybrid Single Particle Lagrangian Integrated Trajectory (HYSPLIT) model,^48^ School Building Archetype (SBA) model using both the Wells-Riley and Monte-Carlo approaches,^44^ the multi-region FATE model of Fauske & Associates,^45^ and dose-response model.^49^ Viruses investigated include SARS-CoV-2,^45,49^ measles with influenza size distributions,^44^ influenza,^42-43,46^ rhinovirus,^46^ PRRSV,^47^ avian influenza,^48^ and virus modelled as gas.^41^ Augenbraun et al^49^ sought to examine SARS-CoV-2 but the relevant infectivity parameters were set to that of influenza. A wide variety of settings were analyzed including a twin aisle airliner cabin,^41^ an office building,^42^ a cruise ship,^43^ a school building,^44^ a detached home,^46^ sow barn,^47^ a farm,^48^ and a quantum physics lab.^49^

From the nine modelling studies, five studies found that increasing filter efficiency associated with decreased virus concentration^41,46^ and decreased risk, including absolute risk and relative risk of infection,^42^ transmission risk,^44^ and infection risk^48^ (see Table 5). One study found that the use of filtration was associated with decreased attack rate.^43^ Two studies found HEPA filtration was associated with decreased risk of SARS-CoV-2 infection^45^ and decreased probability of SARS-CoV-2 infection.^49^ Finally, Janni et al^47^ found that a combination of MERV 8 prefilter and MERV 16 was associated with a lower concentration of PRRSV compared to a combination of MERV 8 prefilter and MERV 14.

Of the nine modelling studies, four evaluated the use of a single filter^43,45,48-49^; all agreed that installing a filter is more beneficial than not installing a filter. Like the experimental studies of Dee et al^36^ and Spronk et al,^37^ and three modelling studies comparing single filters^41-42,46^ agreed that there is a threshold beyond which increasing filter efficiency may not affect the probability of infection. One study using a MERV 8 prefilter was able to model a difference in virus concentration comparing MERV 14 and MERV 16.^47^ Mazumdar and Chen^41^ compared HEPA filters of different efficiencies and determined that improvement was not evident when increasing filter efficiencies past 99.90%. Azimi and Stephens^42^ and Brown et al^46^ specifically found that there were diminishing returns for MERV 13 or higher filters when using influenza, while Dee et al^36^ and Spronk et al^37^ found no difference between MERV 14 and MERV 16 filters for PRRSV. Azimi and Stephens,^42^ using influenza, compared varying MERV filters and a HEPA filter and found that, for filters MERV 13 or greater, the risk reduction was not limited by the filter efficiency; instead by the amount of recirculating air through the HVAC system. Similar to Washam et al,^27^ Azimi and Stephens^42^ considered the cost of filtration. Compared with the cost of an equivalent amount of outdoor air ventilation, Azimi and Stephens^42^ found filtration was the less expensive option for all MERV ratings except MERV 4.

### Risk of Bias

Twelve of the 14 experimental studies had low risk of bias for all three domains: selection bias, information bias, confounding (Table 6). Two experimental studies had low risk of bias for selection bias and confounding but had unclear risk of bias for information bias because of a lack of clarity in the methodological details.^28-29^ Seven of the nine modelling studies had low risk of bias for all three domains: definition, assumption, validation (Table 7). Two modelling studies had low risk of bias for assumption and validation but had unclear risk of bias for definition because of a lack of clarity about the HEPA filter efficiency^45^ and contribution of fresh air.^49^

**Table 6.** Risk of Bias for Experimental Studies.

| **Study** | **Selection Bias** | **Information Bias** | **Confounding\*** |
| --- | --- | --- | --- |
| Washam (1966)<sup>27</sup> | low | low | low |
| Hopkins (1971)<sup>39</sup> | low | low | low |
| Dee (2006)<sup>33</sup> | low | low | low |
| Dee (2009)<sup>34</sup> | low | low | low |
| Pitkin (2009)<sup>35</sup> | low | low | low |
| Malaithao (2009)<sup>28</sup> | low | unclear | low |
| Spronk (2010)<sup>37</sup> | low | low | low |
| Dee (2011)<sup>36</sup> | low | low | low |
| Alonso (2013)<sup>38</sup> | low | low | low |
| Kunkel (2017)<sup>40</sup> | low | low | low |
| Wenke (2017)<sup>30</sup> | low | low | low |
| Bandaly (2019)<sup>29</sup> | low | unclear | low |
| Zhang (2020)<sup>26</sup> | low | low | low |
| Vyskocil (2021)<sup>31</sup> | low | low | low |

\* Confounding assessed for our comparison of interest.

**Table 7.** Risk of Bias for Modelling Studies.

| **Study** | **Definition** | **Assumption** | **Validation** |
| --- | --- | --- | --- |
| Mazumdar (2009)<sup>41</sup> | low | low | low |
| Azimi (2013)<sup>42</sup> | low | low | low |
| Brown (2014)<sup>46</sup> | low | low | low |
| Zheng (2016)<sup>43</sup> | low | low | low |
| Janni (2018)<sup>47</sup> | low | low | low |
| Zhao (2019)<sup>48</sup> | low | low | low |
| Augenbraun (2020)<sup>49</sup> | unclear | low | low |
| Azimi (2020)<sup>44</sup> | low | low | low |
| Kennedy (2020)<sup>45</sup> | unclear | low | low |

## Discussion

While there is substantial literature on the use of filters to remove particles, this review was designed to consider the use of filters to remove viruses. The 23 articles that satisfied our strict criteria revealed several important findings. First, filtration was associated with decreased virus transmission. Second, filters removed viruses from the air. Third, increasing filter efficiency (efficiency of particle removal) was associated with decreased transmission, decreased infection risk, and increased viral filtration efficiency (efficiency of virus removal). Fourth, increasing filter efficiency above MERV 13 was associated with limited benefit in further reduction of virus concentration and infection risk. Fifth, filters with the same filter efficiency rating from different companies had varied performance regarding transmission in animal studies.

### Implications for Research

The modelling studies spanned from 2009 to 2020 and the aerosolized virus studies spanned from 1966 to 2021; some of the more recent studies were motivated by the COVID-19 pandemic. Many viruses were examined; however, it is particularly interesting that there were only two coronavirus studies (both SARS-CoV-2) for filtration, both modelling, and only one aerosolized study that used MS2 as a potential surrogate for SARS-CoV-2 based on size.^26^ Additionally, one of the studies^49^ discussed the implications of filtration in relation to SARS-CoV-2 but used influenza to set relevant infectivity parameters. The limited number of coronavirus (and specifically SARS-CoV-2) studies suggests a potential research gap to be addressed in the future. There is a very rich body of literature of engineering or laboratory studies which focuses on collecting physical metrics expected to affect transmission; these were not considered in this search as our focus was on connecting transmission itself with installed equipment. The trade-off between isolating specific parameters related to filtration physics or analysing more combined links in the chain of transmission is an important consideration and we stress that the literature gaps tend to be in studies which consider systems holistically. This points to a need for interdisciplinary studies which combine knowledge of well controlled filter experiments with less well controlled “real world” situations which inherently capture combined synergies in the transmission chain as well as possible “unknown unknowns” in that chain.

Interestingly, the animal studies only spanned from 1971 to 2013. Furthermore, no field studies examining potential mitigation of virus transmission in humans using filtration were found. This suggests a research opportunity to investigate the use of filtration on mitigating virus transmission in “real-world” built environments occupied by humans, as well as epidemiological studies during disease outbreaks. Taken together, these findings identify an opportunity for more research that investigates HVAC filtration mitigation of human viruses and, specifically, human coronaviruses.

More recent aerosolized virus studies use the ASHRAE Standard 52.2 test duct.^26,31^ This establishes an important level of consistency for future research. The aerosolized virus studies that state an explicit interest in SARS-CoV-2 used the bacteriophage MS2, which has a smaller size than SARS-CoV-2. Future research could explore the ramifications, if any, of this size differential on virus transmission given the size of droplet or droplet nuclei in which the virus itself is suspended.

### Implications for Practice

Some findings are straightforward to address in practice. For example, proper installation is required for a filter to be effective in removing virus and preventing transmission.^39^ Other findings are challenging to address in practice. Two studies documented variations in the performance of similarly rated filters that were purchased from different manufacturers.^36,39^ Variations in performace can occur in filters with the same MERV rating from the same manufacturers, not just different manufacturers. This alone implies that within the broad criteria of MERV ratings, there is enough variation in performance to point to a need for more filter sub-ranges or more stringent testing methods of MERV value.

A practical benchmark emerging from this review was, from the findings of Zhang et al,^26^ that a conservative estimate of the viral filtration efficiency could be approximated by the E1 efficiency used to determine MERV rating in ASHRAE Standard 52.2-2017. Not surprisingly, increasing filter efficiency, whether based on increasing filter efficiency before MERV ratings were established,^39^ increasing MERV rating,^42^ or increasing from MERV to HEPA,^34^ was associated with mitigation of virus transmission. An important practical consideration is that the improvement is limited above the filter efficiency MERV 13. This concept of diminishing returns was documented in both animal studies^36-37^ and modelling studies.^41-42,46^ Spronk et al^37^ and Dee et al^36^ found that both MERV 14 and MERV 16 were associated with no transmission, while Azimi and Stephens^42^ and Brown et al^46^ showed that filters rated MERV 13 or better were associated with limited improvement. One model demonstrated decreased virus concentration with MERV 14 versus MERV 16 but both had MERV 8 prefilters.^47^ While this could suggest an important role of prefilters at higher MERV ratings, the results are difficult to generalize across MERV ratings because only two were modelled. Although one study investigated prefilters, no studies examined filter replacement frequency within the context of viral filtration efficiency.

Diminishing returns are not limited to MERV ratings as Washam et al,^27^ prior to the use of MERV ratings, found that similar efficiency can be achieved using a lower cost option. In addition, the cost increases with increasing MERV rating and HEPA filtration relative to the cost for MERV 13; however, the cost of filtration for these filters remains less than the cost of outdoor air ventilation with the equivalent transmission reduction.^42^ This finding is consistent with current ASHRAE recommendations for reducing airborne infectious aerosol exposure.^12^ Interestingly, ASHRAE indicates that MERV 13 is recommended but MERV 14 is preferred.^13^ The use of MERV 14 is supported by WHO in the context of COVID-19.^14^

Installing or upgrading an air filtration system can sometimes be an expensive intervention. However, illness costs money as well and there is considerable research available showing simply how improved indoor air quality (IAQ) infrastructure investment is profitable when building owners and users interests are aligned^50^. Installing air filtration on large sow farms is estimated to cost $150-200 per sow^51^. Alonso et al^38^ postulates that air filtration like this is quite common now due to the major economic impact of PRRSV on swine production. When considering poultry, Hopkins and Drury^39^ agreed that the additional cost of high efficiency filters would be justified by the investment return of the flock. A single influenza case results in an approximate economic loss of $375 in the United States^52^. Azimi and Stephens^42^ found that upgrading a MERV 7 filter to MERV 13 would only be $17 in labour cost annually in a typical office environment while reducing the number of infections by 1. Not only does air filtration reduce the number of viral infections, it improves the overall air quality by reducing the amount of allergens and asthma triggers in the air. An extra $50 per year for high efficiency air filters in a family residence is seen as a small additional cost against the enormous financial and health burden of asthma^46^.

Some studies have shown that there can be a plateau in effectiveness for filters past MERV 13 in typical situations^42,46^. Mazumdar and Chen^41^ found that a 94% efficiency filter would not be able to protect passengers sitting far away from the infectious source in an airliner cabin and that a 99.9% filter was needed for their application. They go on to state that further increase in filter efficiency might be difficult and its effectiveness not clear. All filters with the same rating are not equal either. Differences in the filter media or frame design across companies and inconsistency in the filter rating systems across countries can contribute to discrepancies in filter efficiency^34^. The source of the raw material of the filter media can also impact filter quality^35^. Zhang et al^26^ found that the viral filtration efficiency (VFE) is generally correlated with the MERV rating but they are not the same. They found that the VFE is always higher than E1, but lower than E2 or E3 efficiencies. This is all to say that choosing the right filter for the right application is important. The location, use case, existing system, budget, and acceptable level of risk should be taken into account when considering air filtration systems^33,36,37^.

When considering scientific testing of filters, the size of the aerosol challenge should be considered very carefully. Many studies have stated that the level of contamination in their experiments was probably much larger than what would be expected in their respective typical situations^27,34,39,40^. This could lead to certain interventions being wrongfully deemed inadequate.

Filtration should not be seen as the only approach to infection mitigation. Filtration should be paired with other infection mitigation strategies such as vaccination^44^, distancing shared space users, and implementing a wait time before different users access the space^49^, and other strict biosecurity measures^30^. Installation of a high efficiency filter increases the pressure drop which can lead to greater energy use if the system has to run more often to service the same volume of air. If installing or upgrading the air filtration system is not possible, Bandaly et al^29^ suggests switching to 100% outdoor air because some viral particles that pass through an inferior filter will still be infectious.

From our review of the literature, it is clear that upgrading or installing high efficiency air filtration is cost effective in the long run and reduces virus transmission. It is recognized that not all spaces are the same and it might not be feasible to implement high efficiency air filtration. If this is the case, then other infection mitigation measures such as UVGI, increasing ventilation rate, or controlling the relative humidity should be considered.

## Conclusion

Evidence clearly demonstrates that filters are effective in removing viruses from the air and reducing the potential for virus transmission and risk of infection. However, experimental and modelling studies indicate diminishing returns above MERV 13, with associated cost implications. Further, above MERV 13, other HVAC features (e.g., amount of recirculating air) may impact risk reduction beyond filtration efficiency. This systematic review identified no field studies or epidemiological investigations of the “real-world” effectiveness of filters in mitigating virus transmission in humans, representing an important gap and priority for future research. Only two modelling studies of SARS-CoV-2 and two experimental studies of MS2 were directly relevant to the current COVID-19 pandemic and recent coronavirus outbreaks.

## Data Availability

All data used in this review come from publicly available indexed journal publications by other authors.

## Acknowledgements

We thank Tara Landry and Alison Henry for conducting the peer review of the search strategies. We thank Samuel Ducholke, Kristen Rumbold, Larry Zhong, and Stella Mathews for their involvement in screening studies for inclusion.

## Notes

**Conflict of Interest:** Dr. Fleck is an unpaid advisor for Pura Air Inc. in Vancouver.

**Funding:** This work is funded by a Canadian Institutes of Health Research (CIHR) Operating Grant: Canadian 2019 Novel Coronavirus (COVID-19) Rapid Research Funding Opportunity [https://webapps.cihr-irsc.gc.ca/decisions/p/project_details.html?applId=422567&lang=en]. Dr. Hartling is supported by a Canada Research Chair in Knowledge Synthesis and Translation. Drs. Fleck and Zhong are supported by NSERC Discovery program.

### Competing Interest Statement

Dr. Fleck is an unpaid advisor for Pura Air Inc. in Vancouver

### Funding Statement

Canadian Institutes of Health Research (CIHR) Operating Grant: Canadian 2019 Novel Coronavirus (COVID-19) Rapid Research Funding Opportunity (Zhong,Hartling & Fleck). Canada Research Chair in Knowledge Synthesis and Translation (Hartling). NSERC Discovery (Zhong and Fleck)

### Author Declarations

Not applicable because this is a systematic review.

